# A neonatal ward strengthening program improves survival for neonates treated with CPAP at district hospitals in Malawi

**DOI:** 10.1101/2021.11.15.21266349

**Authors:** Jennifer Carns, Sara Liaghati-Mobarhan, Aba Asibon, Samuel Ngwala, Elizabeth Molyneux, Maria Oden, Rebecca Richards-Kortum, Kondwani Kawaza, Alfred Chalira, Norman Lufesi

## Abstract

**Introduction:** From 2013-2015, a CPAP quality improvement program (QIP) was implemented to introduce and monitor CPAP usage and outcomes in the neonatal wards at all government district and central hospitals in Malawi. In 2016 the CPAP QIP was extended into healthcare facilities operated by the Christian Health Association of Malawi. Although clinical outcomes improved, ward assessments indicated that many rural sites lacked other essential equipment and a suitable space to adequately treat sick neonates, which likely limited the impact of improved respiratory care. The aim of this study was to determine if a ward strengthening program improved outcomes for neonates treated with CPAP.

**Methods:** To address the needs identified from ward assessments, a ward strengthening program was implemented from 2017-2018 at rural hospitals in Malawi to improve the care of sick neonates. The ward strengthening program included the distribution of a bundle of equipment, supplemental training, and, in some cases, health facility renovations. Survival to discharge was compared for neonates treated with CPAP at 12 rural hospitals for one year before and for one year immediately after implementation of the ward strengthening program.

**Results:** In the year prior to ward strengthening, 189 neonates were treated with CPAP; in the year after, 232 neonates received CPAP. The overall rate of survival for those treated with CPAP improved from 46.6% to 57.3% after ward strengthening (p=0.03). For the subset of neonates with admission weights between 1.00 – 2.49 kg diagnosed with respiratory distress syndrome, survival increased from 39.4% to 60.3% after ward strengthening (p=0.001).

**Conclusion:** A ward strengthening program including the distribution of a bundle of equipment, supplemental training, and some health facility renovations, further improved survival among neonates treated with CPAP at district-level hospitals in Malawi.

## Introduction

Globally, 85% of neonatal deaths are attributed to complications from prematurity and low birthweight, intrapartum-related hypoxia, and infection [1]. Neonates with these complications are particularly susceptible to respiratory distress, thermal instability, infection, jaundice, and hypoglycaemia [2]. Although delivery in a health facility can reduce the risk of neonatal mortality by 29% in low and middle income countries [3], facilities must have the infrastructure, capacity, and resources to treat small and sick neonates for such reductions in mortality to be realized [4,5]. Healthcare facilities must be able to provide neonatal care (service availability) through the presence of essential infrastructure, functioning equipment, supplies, medicine, trained staff, and current care guidelines (service readiness) [5].

Over 91% of deliveries in Malawi occur in a health facility [6]. However, improvements are needed in health facility service availability and service readiness to provide level-2 neonatal care [7], particularly in rural areas where 83% of Malawi’s population live [8] and where 85% of healthcare facilities in Malawi are located [5].

To address limitations in the capacity to treat neonates with respiratory distress, a quality improvement program (QIP) for neonatal care in Malawi was implemented in 2013-2015 to introduce the use of bubble Continuous Positive Airway Pressure (CPAP) at all government district and central hospitals in Malawi [9]. In 2016 the CPAP QIP was extended to 8 healthcare facilities operated by the Christian Health Association of Malawi (CHAM), the largest non-governmental healthcare provider and trainer of healthcare providers in Malawi. Equipment to support respiratory care (CPAP machines, oxygen concentrators, suction machines, pulse oximeters, all necessary disposable supplies, a storage cabinet, and wall job aids) was installed; training on how to use equipment was conducted and supportive supervision and mentorship were provided [10,11]. This was followed by a reduction in neonatal mortality, especially that caused by Respiratory Distress Syndrome (RDS) [9]. However, after conducting detailed needs assessments at each facility, it became increasingly clear that many neonatal units in rural areas lacked other essential equipment and infrastructure, and that further mortality reductions could only be achieved by taking a broader view of the overall challenges experienced by district-level hospitals. Therefore, in 2017-2018, a ward-strengthening program was implemented to provide a bundle of neonatal care services in a dedicated, well equipped clinical space with training and mentorship extended to all relevant hospital technicians and maintenance staff, as well as clinical staff. The aim of this study was to determine if the ward strengthening program improved outcomes for neonates treated with CPAP at these facilities. Here, we describe the process and outcome of this ward strengthening program.

## Methods

### Needs Assessments

In response to the challenges experienced during the CPAP QIP, detailed neonatal ward assessments were done in 2017 at all government district hospitals and CHAM facilities to determine specific challenges and limitations at each site. These assessments documented demand and capacity (number of monthly admissions, number of beds, dimensions of neonatal unit), staffing levels (number of nurses and doctors allocated to the neonatal unit or shared between units), and equipment inventory (functioning and non-functioning equipment allocated to the neonatal unit). Facilities lacking essential equipment and/or suitable space for a neonatal unit were selected for inclusion in a ward strengthening program.

We also assessed neonatal training programs at fifteen institutions that train nurses and clinicians to ensure sustainability in the transfer of knowledge and skills. The assessment showed that, despite the presence of neonatal care concepts in training curricula at all institutions, there were gaps in teaching these concepts, particularly due to poorly furnished skills labs. All fifteen training institutions were included in a program to strengthen skills labs for neonatal care.

### Ward Strengthening Program

From 2017-2018, a ward strengthening program was implemented at 12 government district hospitals and CHAM facilities to improve the care of sick neonates. Ward strengthening activities included the distribution of a bundle of equipment (Table 1), supplemental training support, and, in some cases, health facility renovations (Table 2).

**Table 1.**
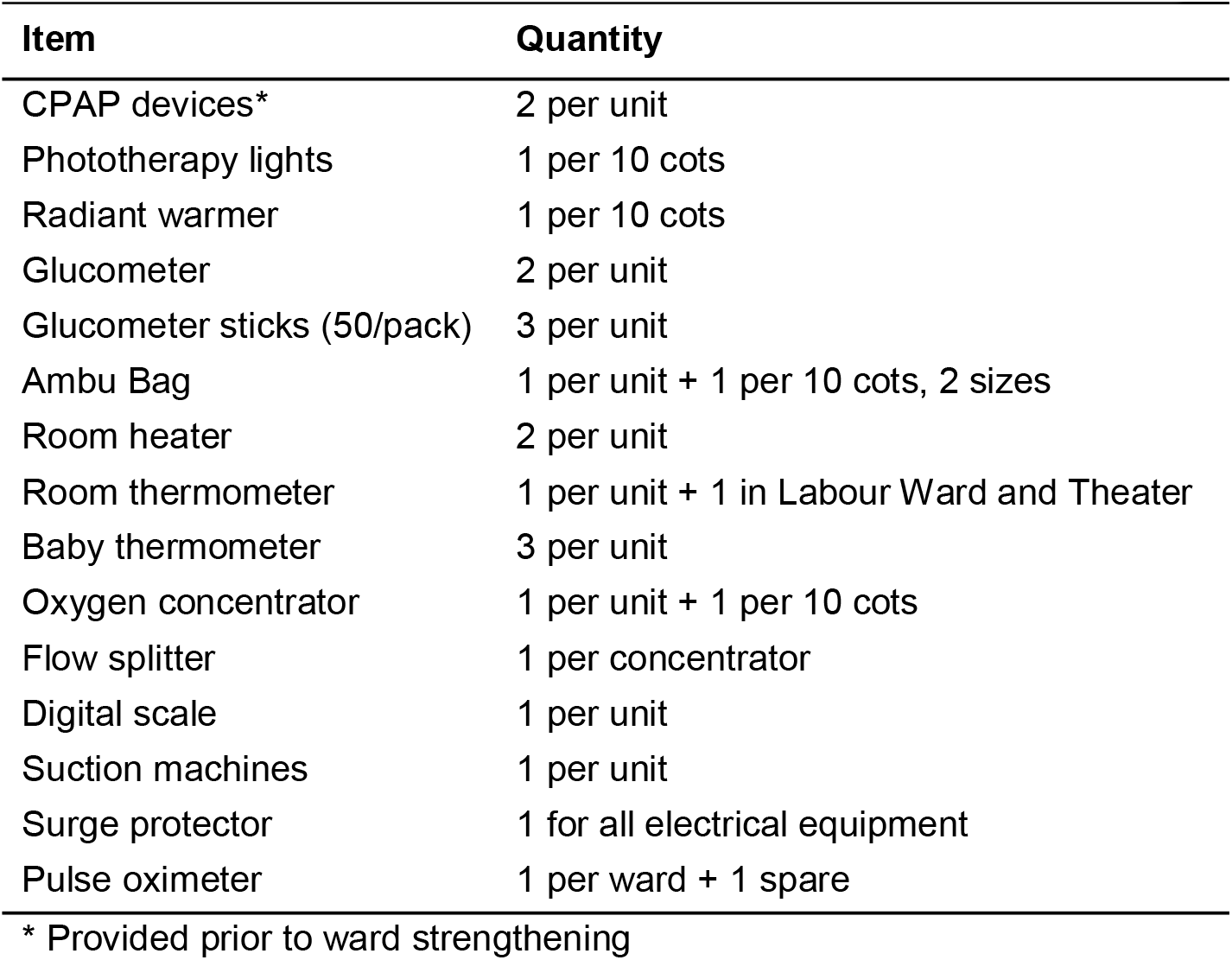
Equipment bundle and quantity by number of beds.

**Table 2.**
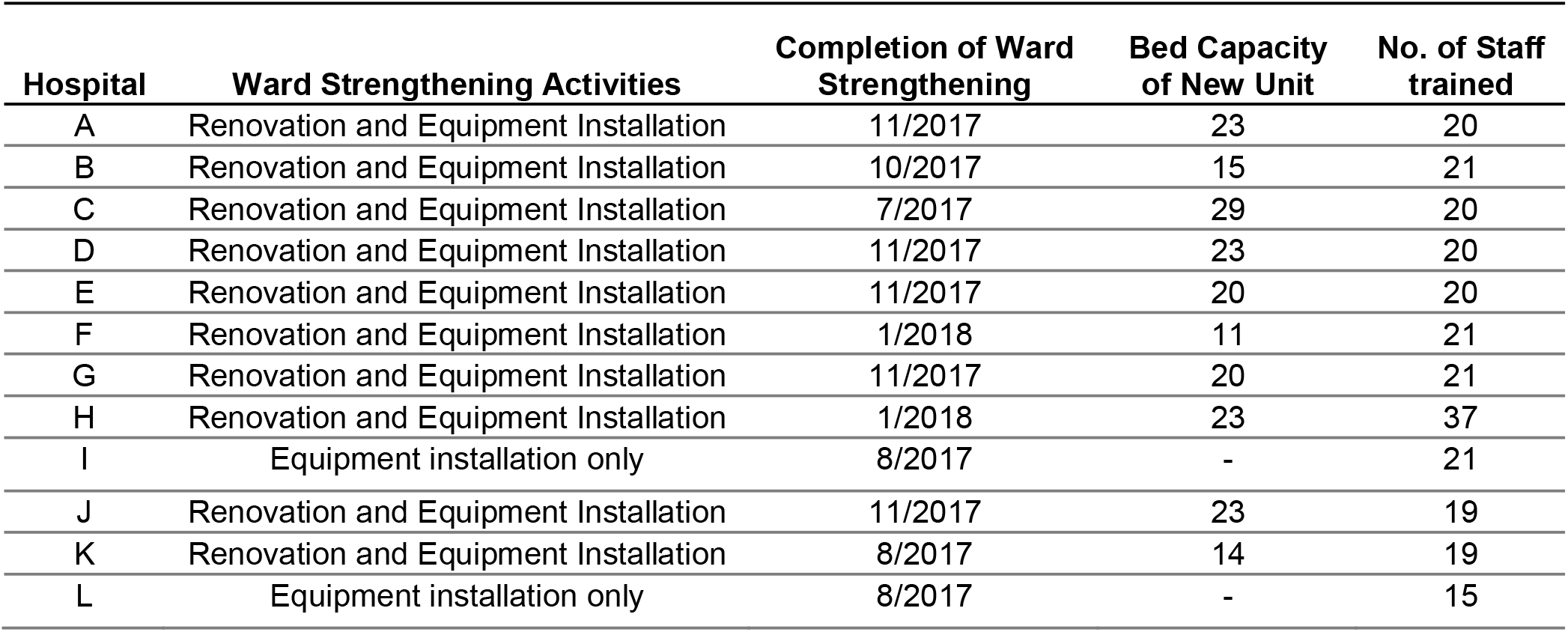
Summary of Renovations by Facility.

Ten of the 12 facilities lacked a separate or suitable neonatal unit. At these sites, the facility management team worked with the Ministry of Health (MOH) to identify a room or space for renovation, quantify material needs for the renovation, obtain quotations for the materials, and develop a renovation work plan. Renovations varied by facility, but generally included the expansion or partitioning of an existing space, plumbing installation and repairs, installation of lighting and electrical sockets, roofing and ceiling installation and repairs, and painting. For nine of the ten sites where renovations were completed, renovation costs ranged from $1,300 - $8,600 USD and were covered by project funds. At the remaining site, the cost of construction of a new ward ($38k USD) was supplemented by private donor funds.

At each site, equipment was distributed according to the bed capacity of the unit (Table 1); for a unit with 20 cots the bundle of equipment was approximately $14k USD. Before equipment installation, clinical staff and biomedical technicians completed half-day user training courses to ensure familiarity with the operation, troubleshooting and maintenance of the equipment. These trainings were conducted with the Ministry of Health Physical Asset Management (PAM) units to allow biomedical technicians to receive training as well as assist with the instruction and installation process at each site. Costs associated with trainings included travel costs for the instructors and per diems for the trainees. The number of staff trained at each facility is summarized in Table 2.

In collaboration with the Paediatric and Child Health Association of Malawi (PACHA), training of the nurses and clinicians was delivered at the facilities using the Care of the Infant and Newborn (COIN) course. The COIN course, which is used nationally, focuses on diagnosis and triage of sick neonates, care at birth for all neonates, neonatal resuscitation, routine postnatal care of the neonate, emergency assessment and treatment of neonates, and identification of neonates in need of referral and safe transport [12].

After the completion of the ward strengthening program, follow-up ward assessments were completed at each healthcare facility. The follow-up ward assessments were compared to those conducted prior to the ward strengthening program to evaluate changes in demand, capacity, and staffing levels.

### Program to Strengthen Skills Labs for Neonatal Care

Fifteen health training institutions received support to establish skills labs for neonatal care, including equipment and teaching materials to facilitate hands-on teaching of essential skills in the care of neonates. Materials provided included training models, oxygen concentrators, glucometers, CPAP machines, suction machines, phototherapy lights, and teaching aids. Academic staff from these institutions also received training to encourage use of these resources within their institutions.

### Study Setting

The impact of the ward strengthening program was evaluated at 12 district and CHAM hospitals located in rural areas (Table 2) where CPAP was being used to treat respiratory distress for one year before, and one year after the implementation of the ward strengthening program. All neonates with known outcomes (i.e. died or survived to discharge) treated with CPAP for respiratory distress at each facility were included in this analysis. Clinical diagnoses were made in accordance with national guidelines for newborn care in Malawi [12]. Because gestational age is often unknown in our setting, a simple, validated algorithm using a combination of vital signs, tone and birth weight was implemented to assist nurses in determining the need for CPAP [13,14]. As previously described [9], patient details including dates of birth and admission, admission weight, discharge diagnosis, and outcome were collected from Acute Respiratory Illness (ARI) forms for each neonate admitted with respiratory illness. An on-site ARI coordinator ensured completion of forms for every qualifying patient. De-identified ARI forms were scanned monthly, and quarterly chart audits were conducted by the MOH ARI team to ensure completion of ARI forms for all patients. Patient data and survival data were compared for neonates treated before and after ward strengthening. Neonates with unknown outcomes, including those that left against medical advice or were transferred to another facility, were excluded from analysis. Rate of survival was defined as the fraction of eligible neonates with known outcomes who survived to discharge.

### Statistical Analysis

Differences in transfer rates and survival before and after ward strengthening were compared using a two-sided Fisher’s exact test. Differences between continuous variables (i.e., weight and age on admission) were assessed using a two-sided t-test for equality of means (unequal variances assumed). Results were considered significant at the 5% level. Data were analyzed in Excel.

### Ethics statement

The protocol was approved by the National Health Sciences Research Committee (NHSRC #1180) of Malawi and the Institutional Review Board of Rice University (13-102X). De-identified patient information was collected from standard Ministry of Health Acute Respiratory Illness forms.

## Results

Table 3 summarizes the patient data, outcomes, and survival rates for neonates treated with CPAP one year before and after ward strengthening. A total of 207 neonates were treated with CPAP before ward strengthening, and 254 were treated after ward strengthening. Among neonates with known outcomes, 189 were treated with CPAP before ward strengthening, and 232 were treated with CPAP after ward strengthening. There was no significant difference in the number of neonates who transferred out of these facilities before and after ward strengthening, and there was no significant difference in the number of neonates who left against medical advice before and after ward strengthening. There were no significant differences in the rates of diagnosis of birth asphyxia, RDS, pneumonia, and meconium aspiration before and after ward strengthening. The percentage of neonates with known outcomes and a final diagnosis of sepsis significantly decreased after ward strengthening (12.2% vs 4.7%, p=0.01).

**Table 3:** Patient data for neonates treated with CPAP for one year before and after ward strengthening.

|  | Before Ward<br>Strengthening | After Ward<br>Strengthening | p-value |
| --- | --- | --- | --- |
| **Number of Study Participants** | 207 | 254 |  |
| **Outcome** |  |  |  |
| Died | 48.8% | 39.0% | **0.038** |
| Discharged | 42.5% | 52.4% | **0.039** |
| Transferred | 5.3% | 3.5% | 0.368 |
| Left AMA | 3.4% | 3.9% | 0.809 |
| Unknown | 0.0% | 1.2% | 0.256 |
| **Number of Neonates With**<br>**Known Outcome**<br>**(Died/Discharged)** | 189 | 232 |  |
| **Outcome** |  |  |  |
| Died | 53.4% | 42.67% | **0.031** |
| Discharged | 46.6% | 57.33% | **0.031** |
| **Diagnosis\*** |  |  |  |
| Birth Asphyxia | 15.9% | 9.9% | 0.077 |
| RDS | 68.8% | 66.8% | 0.677 |
| Pneumonia | 9.5% | 9.1% | 0.868 |
| Meconium Aspiration | 5.3% | 4.7% | 0.825 |
| Sepsis | 12.2% | 4.7% | **0.007** |
| No Diagnosis | 1.6% | 1.7% | 1.000 |
| **Admission Weight** |  |  |  |
| <1.00 kg | 5.8% | 3.45% | 0.345 |
| 1.00-1.49 kg | 37.0% | 40.5% | 0.483 |
| 1.50-1.99 kg | 23.3% | 23.3% | 1.000 |
| 2.00-2.49 kg | 9.0% | 11.2% | 0.519 |
| 2.50-4.00 kg | 21.7% | 16.8% | 0.214 |
| >4.00 kg | 1.1% | 1.7% | 0.695 |
| Unknown | 2.1% | 3.0% | 1.000 |
| **Admission Temperature** |  |  |  |
| <32.0 °C | 0.0% | 0.86% | 0.504 |
| 32.0-34.4 °C | 22.8% | 24.6% | 0.730 |
| 34.5-35.4 °C | 16.9% | 23.7% | 0.092 |
| 35.5-36.4 °C | 27.5% | 21.6% | 0.171 |
| 36.5-37.5 °C | 13.8% | 13.4% | 1.000 |
| >37.5 °C | 7.9% | 7.8% | 1.000 |
| Unknown | 11.1% | 8.2% | 0.321 |

\*Some neonates received multiple diagnoses

Following implementation of the ward strengthening program, there was a significant increase in survival for neonates treated with CPAP (Fig 1A), for the subset of neonates treated with CPAP weighing between 1.00 – 2.49 kg on admission and diagnosed with respiratory distress syndrome (RDS) (Fig 1B), as well as for the subset of neonates treated with CPAP weighing between 1.00 – 1.49 kg on admission and diagnosed with respiratory distress syndrome (RDS) (Fig 1C).

**Fig 1.**
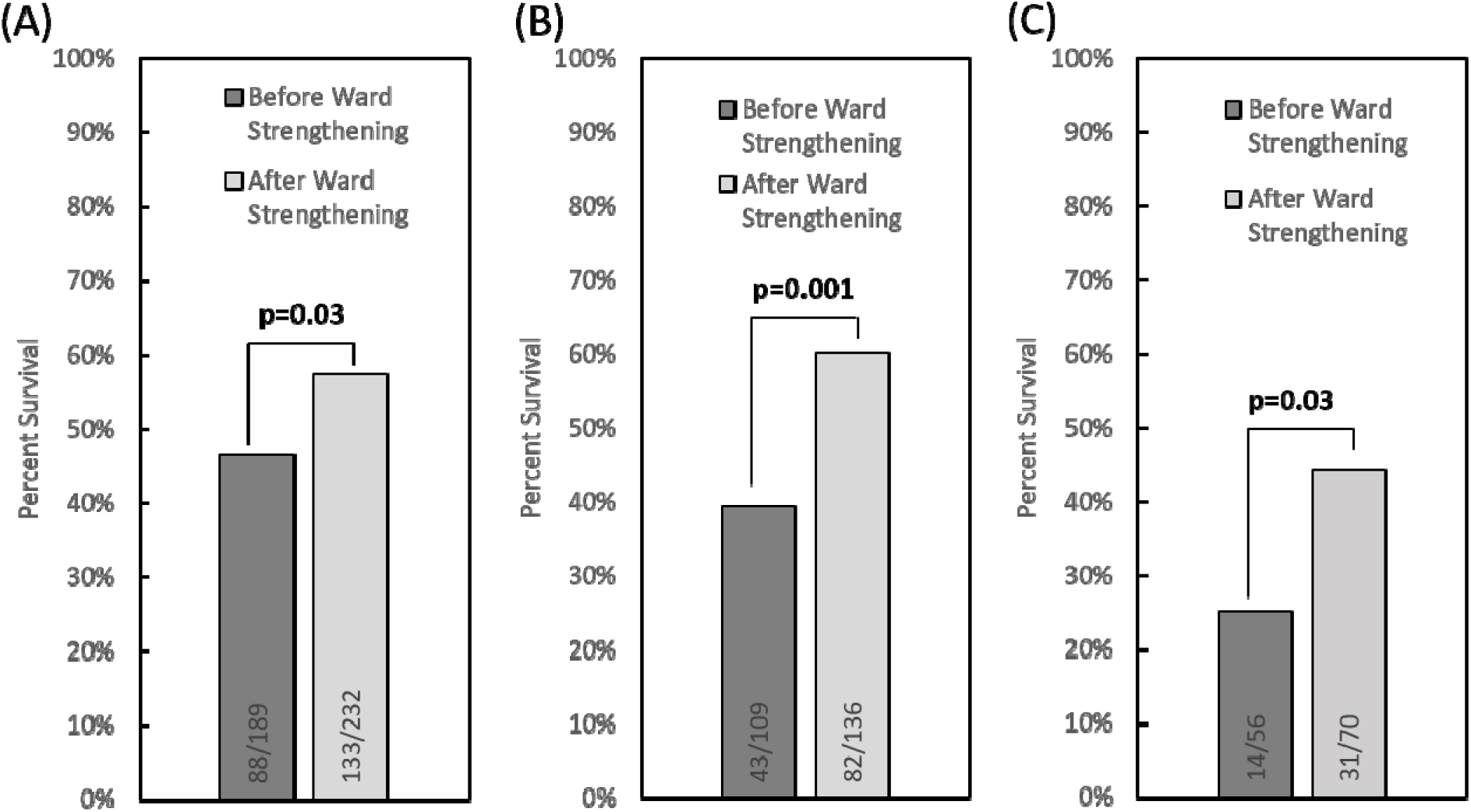
(A) Outcomes for all neonates admitted with respiratory distress and treated with CPAP for one year before and after ward strengthening. There was a significant increase in survival after ward strengthening (46.6% and 57.3%, respectively, p=0.03). (B) Outcomes for the subset of neonates treated with CPAP weighing between 1.00 – 2.49 kg on admission and diagnosed with respiratory distress syndrome (RDS). There was a significant increase in survival after ward strengthening (39.4% and 60.3%, respectively, p=0.001). (C) Outcomes for the subset of neonates treated with CPAP weighing between 1.00 – 1.49 kg on admission and diagnosed with respiratory distress syndrome (RDS). There was a significant increase in survival after ward strengthening (25.0% and 44.3%, respectively, p=0.03).

Before ward strengthening, 46.6% of neonates treated with CPAP survived to discharge; the rate of survival improved to 57.3% after ward strengthening (p=0.03). Survival was also compared in the subset of low birth weight neonates diagnosed with RDS and weighing 1.00 – 2.49 kg and 1.00 – 1.49 kg on admission; in these groups, survival increased from 39.4% before ward strengthening to 60.3% after ward strengthening (p=0.001), and from 25.0% before ward strengthening to 44.3% after ward strengthening (p=0.03).

The admission weights, admission temperatures, and age on admission for neonates treated before and after ward strengthening are compared in Table 4. Results are shown for all neonates treated with CPAP, and for the subset with admission weights between 1.00 – 2.49 kg and diagnosed with RDS. There were no significant differences in the distribution of patient variables before and after ward strengthening, with the exception of a decrease in mean admission temperature for neonates weighing between 1.00 – 2.49 kg with a diagnosis of RDS (p = 0.01).

**Table 4:** Comparison of average weights, temperatures, and age on admission for neonates treated one year before and after ward strengthening.

|  | All Admission Weights |  |  | Admission Weights 1-2.49 kg,<br>Diagnosed with RDS |  |  |
| --- | --- | --- | --- | --- | --- | --- |
|  | Before Ward<br>Strengthening | After Ward<br>Strengthening | p-value | Before Ward<br>Strengthening | After Ward<br>Strengthening | p-value |
| Average<br>Admission<br>Weight (kg) | 1.84 +/- 0.81 | 1.84 +/- 0.96 | 0.99 | 1.48 +/-0.34 | 1.48 +/-0.34 | 0.87 |
| Average<br>Admission<br>Temperature<br>(° C) | 35.5 +/- 1.6 | 35.4 +/- 1.6 | 0.48 | 35.4 +/- 1.5 | 34.9 +/- 1.3 | **0.01** |
| Average Age<br>on Admission<br>(day) | 2.0 +/- 5.1 | 2.1 +/- 5.9 | 0.75 | 0.7 +/- 3.1 | 0.4 +/- 2.1 | 0.48 |

Table 5 compares observations from the initial ward assessments completed in 2017 to the follow-up ward assessments completed in 2018. These assessments indicated that admissions increased and staffing levels improved following ward strengthening. Overall, average nursery admissions at the 12 facilities included in this analysis increased from 39.2 neonates per month in 2017 to 74.9 neonates per month in 2018. Additionally, the total number of nurses allocated specifically for neonatal care across these facilities increased for both the day and night shifts. Overall, the median number of nurses allocated specifically to care for neonates during the day increased from 1 to 1.5, and the median number of nurses allocated to neonatal care overnight increased from 0.5 to 1. The number of hospitals with no nurses allocated specifically to care for neonates during the day remained the same before and after ward strengthening (3 hospitals); the number of hospitals with no nurses allocated specifically to care for neonates during the night decreased from 6 to 3. At the conclusion of this program, at least one clinician was assigned to every neonatal ward included in this analysis.

**Table 5:**
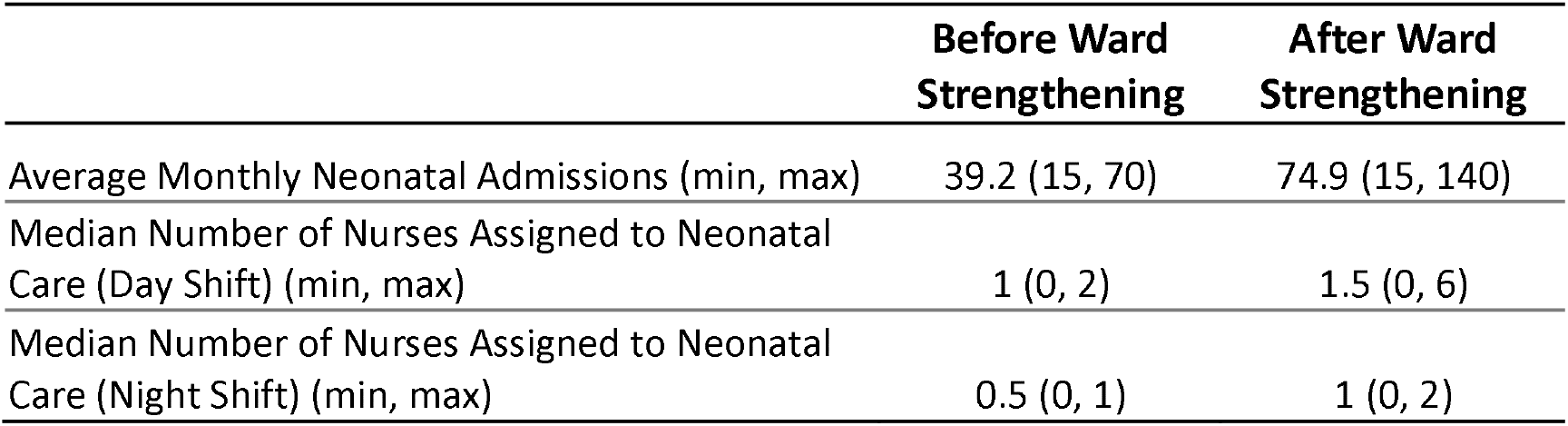
Comparison of ward assessments for average monthly admissions and nurse allocations before and after ward strengthening.

## Discussion

We previously reported that implementation of a nurse-led CPAP QIP to treat neonates with respiratory illness in all government district and central hospitals in Malawi led to a significant increase in survival, particularly among neonates of low birth weight [9]. Following the CPAP QIP, survival for neonates with admission weights of 1.00 – 2.49 kg and diagnosed with RDS increased from 39.8% to 48.3% after the implementation of CPAP (p=0.042) [9]. However, improvements in survival fell short of those experienced during a previous quasi-randomized efficacy study of CPAP vs standard nasal oxygen in the neonatal unit of the referral hospital of southern Malawi, where neonates with admission weights of 1.00 – 2.49 kg and diagnosed with RDS experienced survival rates of 64.6% when treated with CPAP [15].

During mentorship and supervisory visits to rural and district hospitals, many challenges were identified that likely adversely affected overall survival as well as limited the impact of improved respiratory care. Despite provision of supportive respiratory equipment, training, and peer mentorship along with the CPAP devices, additional equipment, training, and space were necessary to allow more comprehensive care of vulnerable neonates, who often have other complications in addition to respiratory distress. After the ward strengthening program was introduced, survival rates for neonates weighing 1.00-2.49 kg with a diagnosis of RDS improved from 39.4% to 60.3%, approaching the 64.6% survival previously seen in a referral hospital [15].

To identify facility-level barriers to improved neonatal care, a needs assessment was undertaken in each hospital by members of their own administrative, clinical and technical/maintenance staff, together with MOH representatives. Together, MOH representatives and research staff determined which parameters to evaluate; only parameters that directly impacted the quality of newborn care were prioritized to avoid burdening hospital staff and prevent disruption of work flow during the assessments. Since hospital staff were familiar with the parameters being evaluated, assessments were completed as part of routine visits to the facilities and no intensive training was required. Building modifications and equipment and staffing needs were thus directed by local requirements. Importantly, this gave ownership and investment to local hospital staff and in itself strengthened interdepartmental relationships.

The most common requirement was that of space. Ten of the twelve facilities did not have a separate neonatal ward; if a separate area was available, it was too small or otherwise unsuitable. The absence of a neonatal ward is common in hospitals in many countries [16,17]. As part of the Every Newborn Action Plan process, an analysis of 12 countries in Africa and Asia with the highest burden of neonatal mortality showed that five countries reported limited or no dedicated neonatal space in healthcare facilities [16]. A recent qualitative assessment of health facilities in three regions of Ethiopia from December 2017 to February 2018 found the lack of space for neonates in need of special care across all levels of the health care system [17]. This assessment found that neonates were sometimes relocated to adult and surgical wards due to the lack of space [17]. A dedicated space is essential for the provision of infection control and temperature control for small and sick neonates. Following the implementation of the ward strengthening program presented here, the proportion of neonates receiving CPAP who were diagnosed with sepsis decreased from 12.2% to 4.7%; further investigation is needed to determine if this decrease was a result of improved diagnoses or improved infection control.

The care of sick neonates also requires specialized training and skills. Neonatal nursing training programs are rare in low-income countries [16], and a high turnover of trained healthcare providers is a problem in many settings [9,17]. We tried to address these barriers by conducting trainings not only at health facilities where equipment was installed, but also at 15 nursing schools and clinical officer training institutions. Most of the training institutions did not have the basic equipment necessary for practical instruction in neonatal care. Strengthening existing pre-service training programs is necessary to address the high staff turnover due to rotations and attrition.

We note that improved space, equipment, and training had important additional beneficial effects. Previously, sick and healthy neonates often remained with their mother in the postnatal ward, but the creation of a separate, well-equipped ward for sick neonates meant that staff were allocated specifically for their care; overall, nurse allocations to the neonatal wards increased and at least one clinician was assigned to the neonatal ward in each facility.

As this observational study involved neonates undergoing routine care in a low resource setting, there were some unavoidable limitations. The primary outcome of this study was survival to discharge as follow-up after discharge was not feasible. Few if any neonates had a chest x-ray to diagnose RDS, and gestational age as determined by a first trimester ultrasound was rarely available. Consequently, RDS was diagnosed clinically with the aid of an algorithm to help identify neonates who might benefit the most from CPAP. Although retinopathy of prematurity was addressed during trainings [11], we did not monitor for it during this study. However, in a review of reports from LMICs on the use of CPAP, two studies that looked for retinopathy of prematurity did not find it [18], and we did not encounter it in the early phases of our program [15]. Adverse events such as pneumothoraces and nasal trauma were also not tracked as part of this study, but anecdotally, they were uncommon and limited to nasal bleeds or nasal hyperemia.

### Learning from our experience

When reflecting on our program, it is clear that efforts to improve the quality of neonatal care must be built on a solid foundation of good basic clinical care that is based on a holistic approach to the needs of a neonate. Improvement in one aspect of need (such as respiratory support) will falter without the provision of warmth, nutrition, infection control and family-centered care given by trained staff in a dedicated space. A needs assessment is essential to identify gaps in overall care so that these can be addressed with an appropriate bundle of interventions to improve care for all infants in the ward. Provision of materials is insufficient without training, supervision, and mentoring to ensure that equipment is used properly and protocols followed correctly.

Some challenges are beyond the jurisdiction of hospital management to solve: an unreliable power system and irregular water supply are common and affect many of the facilities included in this program. While power and water outages were not systematically tracked during this audit, power outages were noted on the charts of 16.4% of neonates treated before, and 15.9% treated after ward strengthening. Furthermore, availability of essential commodities and consumables is a challenge if hospitals lack the resources or ability to procure them.

It is encouraging to see that the physical changes made to the neonatal ward areas led to improved staff allocation to work in them. In initial discussions about possible space renovations, MOH representatives strongly advocated that each facility dedicate staff solely to planned new units to improve quality of care. Based on our observations, better staffing, more training and mentorship have improved morale, and by planning and creating the changes together, the hospital departments are more interactive. All of these factors have played a part in reducing neonatal mortality.

## Conclusion

To improve neonatal outcomes, healthcare facilities need the infrastructure, capacity, and resources to treat small and sick neonates. This means having a family-centred approach to care encompassing the provision of warmth, nutrition and infection control given by trained staff in a dedicated space with functioning equipment. Providing CPAP within such a framework significantly improved survival for neonates with respiratory distress. These improvements in service availability lead to improved staff morale, increased the number of dedicated neonatal staff, and most importantly, improved outcomes for admitted sick and small neonates treated with CPAP.

## Data Availability

All data produced in the present study are available upon reasonable request to the authors

## Acknowledgments

We would like to acknowledge PACHA (Paediatric and Child Health Association) for their leadership in the development and provision of the COIN training course. We would also like to acknowledge all of the staff of the CPAP Project for their contributions to data collection, monitoring, and training associated with the quality improvement initiative, as well as our CPAP trainers and mentors for leading CPAP training sessions and providing mentorship and supportive supervision within the hospitals.

## Contributors

NL, AA, KK, RRK, MO, EM, and AA designed the study. JC, SLM, SN, and AA contributed to data collection and/or data and statistical analysis. JC, SLM, AA, and EM prepared the first draft of the manuscript. All authors contributed to critical interpretation of the results and development of the manuscript and approved the final version.

## Funding

This work is made possible through the generous support of ELMA Philanthropies through a grant to the University of Malawi College of Medicine (https://www.elmaphilanthropies.org/, Grant No. 16-F0007). This manuscript is also made possible through the generous support of the Saving Lives at Birth (https://www.usaid.gov/global-health/health-areas/maternal-and-child-health/projects/saving-lives-birtha-grand-challenge) partners: The United States Agency for International Development (USAID), the Government of Norway, the Bill & Melinda Gates Foundation, Grand Challenges Canada, and the UKAID (Grant No. AID-OAA-A-13-00014). It was prepared by Rice University and does not necessarily reflect the views of the Saving Lives at Birth partners. Additionally, this work was supported by the John D. and Catherine T. MacArthur Foundation (https://www.macfound.org, Grant No. 17-1709-152484) the Bill & Melinda Gates Foundation (https://www.gatesfoundation.org, Grant No. OPP1193531), ELMA Philanthropies (https://www.elmaphilanthropies.org, Grant No. 19-F0012), The Children’s Investment Fund Foundation UK (https://ciff.org, Grant No. R-1810-03159), The Lemelson Foundation (https://www.lemelson.org, Grant No. 18-01458), and the Ting Tsung and Wei Fong Chao Foundation under agreements to William Marsh Rice University.

## Notes

### Competing Interest Statement

Drs. Oden, Richards-Kortum, and Molyneux are inventors on a patent for CPAP that has been licensed to 3SD at zero percent royalty in GAVI eligible countries; all royalties have been donated to Rice University to support global health research and education.

### Funding Statement

This work is made possible through the generous support of ELMA Philanthropies through grant to the University of Malawi College of Medicine. This manuscript is also made possible through the generous support of the Saving Lives at Birth partners: The United States Agency for International Development (USAID), the Government of Norway, the Bill & Melinda Gates Foundation, Grand Challenges Canada, and the UKAID. It was prepared by Rice University and does not necessarily reflect the views of the Saving Lives at Birth partners. Additionally, this work was supported by the John D. and Catherine T. MacArthur Foundation, the Bill & Melinda Gates Foundation, ELMA Philanthropies, The Children's Investment Fund Foundation UK, The Lemelson Foundation, and the Ting Tsung and Wei Fong Chao Foundation under agreements to William Marsh Rice University.

### Author Declarations

The protocol was approved by the National Health Sciences Research Committee (NHSRC #1180) of Malawi and the Institutional Review Board of Rice University (13-102X).

